# Integrated germline and somatic molecular profiling to detect cancer predisposition has a high clinical impact in poor-prognosis paediatric cancer

**DOI:** 10.1101/2024.08.08.24311493

**Authors:** Noemi A Fuentes-Bolanos, Eliza Courtney, Chelsea Mayoh, Meera Warby, Loretta M S Lau, Marie Wong-Erasmus, Dong-Anh Khuong-Quang, Paulette Barahona, Bhavna Padhye, Sam El-Kamand, Sheena Nunag, Pamela Ajuyah, Alexandra Sherstyuk, Ann-Kristin Altekoester, Ashleigh Sullivan, Nicola Poplawski, Catherine Kiraly-Borri, Sarah O’Sullivan, Helen Marfan, Rozanna Alli, Lisette Curnow, Kanika Bhatia, Antoinette Anazodo, Toby N Trahair, Marion Mateos, Jordan R. Hansford, Hetal Dholaria, Sarah Josephi-Taylor, Andrew S Moore, Wayne Nicholls, Nicholas G Gottardo, Peter Downie, Seong-Lin Khaw, Heather Tapp, Geoffrey McCowage, Luciano Dalla-Pozza, Frank Alvaro, Paul J Wood, Vanessa Tyrrell, Michelle Haber, Mark J Cowley, Paul G Ekert, Glenn M Marshall, Judy Kirk, Katherine Tucker, Mark Pinese, David S Ziegler

## Abstract

Germline predisposition has a significant role in paediatric cancer. However, the optimal approach to identifying cancer-causing germline pathogenic variants (GPV) in children, and even the prevalence of GPV among children with cancer, remain unclear. Here we report our findings from a comprehensive survey of GPV in 496 children with poor-prognosis cancer. By integrating tumour and germline molecular profiling we identified GPV in 15.5% of patients, 48.1% of whom had not met clinical genetic testing criteria. Although the cancer type was outside the recognised phenotypic spectrum for 43.7% of reported GPV, 63.2% of these were clinically actionable for cancer risk. Integrated germline-tumour analysis increased the GPV detection rate by 8.5%, and informed germline interpretation in 14.3% of patients with GPV, highlighting the value of integrated analyses. Our findings establish the benefit of broad integrated tumour-germline screening, over phenotype-guided testing, to detect GPV in children with poor prognosis cancers.

## Introduction

Germline cancer predisposition is frequently observed in paediatric cancer patients^1–9^. However, much remains unknown about the prevalence and types of germline variants, their clinical correlates, and relevance to tumour development. In particular, the lack of systematic genetic surveys of children, adolescents, and young adults (C-AYA) diagnosed with cancer has resulted in a knowledge gap limiting the application of genomic medicine to manage cancer risk in the young.

Estimates of the prevalence of germline pathogenic or likely pathogenic variants (GPV) in cancer predisposition genes (CPG) in C-AYA populations have ranged from 8% to as high as 16% ^1–11^ with variance likely due to differences in cohort composition, analytical methodology, and variant classification criteria. This variability in prevalence estimates is clinically important, as accurate prevalence estimates directly inform optimal genetic testing strategies. Definitive estimates of the prevalence of germline cancer risk require consideration of both tumour and germline molecular features ^12^. To our knowledge, however, no cohort study has undertaken such a multi-modal molecular analysis of GPV in C-AYA, leading to uncertainty in prevalence and clinical correlates of cancer risk, negatively impacting patient management.

The optimal approach to identifying GPV in clinical care remains unclear. Prior studies to assess the clinical impact of GPV detection in paediatric cancer have predominantly utilised germline-only whole-genome sequencing (WGS), or tumour sequencing paired with germline whole-exome^8,13^ or panel-based sequencing^7^. The feasibility, clinical utility, and family impact of WGS based screening of GPV in paediatric cancer patients has not yet been evaluated, and it remains unclear whether a WGS-based GPV screening approach should be implemented in routine clinical care.

To address this knowledge gap and evaluate the translation of precision medicine to the cancer genetics clinic, we report the first comprehensive multi-modal survey of GPV in C-AYA with poor prognosis cancers. Leveraging the Zero Childhood Cancer Program’s PRecISion Medicine for Children with Cancer (PRISM) study^9^, we applied current CPG curation guidelines to tumour and germline WGS, tumour RNA sequencing (RNA-Seq), and methylation analysis to definitively assess the prevalence of reportable GPV. We correlated our germline findings to clinical variables and tumour features to evaluate the impact in the interpretation of germline variants. In addition, we assessed the effect of returning these germline cancer risk findings as part of clinical care at a national level.

## Results

### Genetic cancer predisposition is common in poor prognosis paediatric cancer

To evaluate the impact of rapid germline WGS integrated with tumour analysis, we identified a primary cohort of 496 participants consecutively enrolled in the ZERO Childhood Cancer PRISM study from 2018 to 2020^9^. Patients were enrolled on PRISM based on having a poor prognosis cancer (estimated survival <30%) diagnosed by age 21 years, representing all major paediatric cancer categories (Table 1). Patients underwent paired germline and tumour WGS and tumour RNA-Seq, followed by expert curation, and national multidisciplinary tumour board (MTB) discussion ^9^. Findings from the first 247 patients, including 40 patients (16.2%) with GPV,^9^ and precision-guided treatment outcomes of the first 384 patients enrolled in this precision medicine program, were previously reported^14^.

**Table 1.** Characteristics of the 496 individuals in the primary study cohort.

|  | Participants<br>(% of total cohort) | GPV in a CPG<br>(% of subgroup) | *p* value |
| --- | --- | --- | --- |
| **Sex** |  |  | 0.71 (0.71) |
| Female | 231 (46.6%) | 34/231 (14.7%) |  |
| Male | 265 (53.4%) | 43/265 (16.2%) |  |
| **Age at diagnosis (yrs)** | 8 (3-13) | 8 (2-10) | 0.21 (0.10) |
| **Cancer Category<sup>#</sup>** |  |  | 0.081 (0.027) |
| CNS tumours | 184 (37.1%) | 27/184 (14.7%) |  |
| Sarcomas | 131 (26.4%) | 13/131 (9.9%) |  |
| Other solid tumours | 97 (19.6%) | 24/97 (24.7%) |  |
| Haematological malignancies | 84 (16.9%) | 13/84 (15.5%) |  |
| **Family history suggestive of cancer risk\*** |  |  | 0.031 (0.0076) |
| Yes | 27 (11.2%) | 11/27 (40.8%) |  |
| No | 214 (88.8%) | 36/214 (16.8%) |  |

Abbreviations: GPV, germline pathogenic/likely pathogenic variant/s; CPG, cancer predisposition gene; CNS, central nervous system.
\*Cases with family history not available were excluded from the test. #ICC category 3<sup>rd</sup> edition. Categorical variables are reported as counts with frequency, and continuous variables as medians with interquartile range. *p* values are from Fisher's exact tests (categorical variables), or Wilcoxon signed rank tests (continued) and are reported as family-wise error rate corrected *p* values (adjusting for four tests by Holm's step-down procedure), with raw *p* values in parentheses.

We identified and reported to the treating clinician 85 GPV in 77/496 patients (15.5%), with five patients harbouring GPV in more than one CPG ^15^(Supplementary Fig. 1) Family history information was requested from the treating oncologist at enrolment. Reportable GPV were significantly more common in patients with a family history suggestive of cancer risk (40.7% vs 16.8%, Fisher’s exact p = 0.031, Table 1), the rate of GPV was significantly higher in patients who had family history assessed than in patients for whom family history was unavailable or inconclusive (19.5% vs. 11.8%, Fisher’s exact p = 0.019), suggesting that family history documentation may be influenced by the pre-test suspicion of genetic cancer risk. Importantly, 76.6% of patients with a GPV had no relevant family history. No other significant clinical variables (cancer category, participant sex, and age at diagnosis) were significantly associated with the presence of a GPV (*p* > 0.08 following multiple test correction, Table 1). However, well established associations for specific cancer subcategories were evident, for example 4/7 (57.1%) patients with adrenocortical carcinoma had germline *TP53* GPV.

To further characterise the GPV, we investigated the spectrum of cancer risk variants, both at the gene and pathway level. The GPV were reported across 32 genes, with the highest frequency in *TP53*, *CHEK2,* and *NF1* (Fig. 1b). A strong association was observed between the biological pathway linked to the GPV and cancer type (Fig. 1c, Fisher’s exact test p < 0.001), most notably for GPV in the mismatch repair pathway (MMR), which were exclusively observed in patients with central nervous system (CNS) tumours (Fig. 1c). Overall, GPVs were most prevalent in genes associated with DNA repair and maintenance.

**Figure 1.**
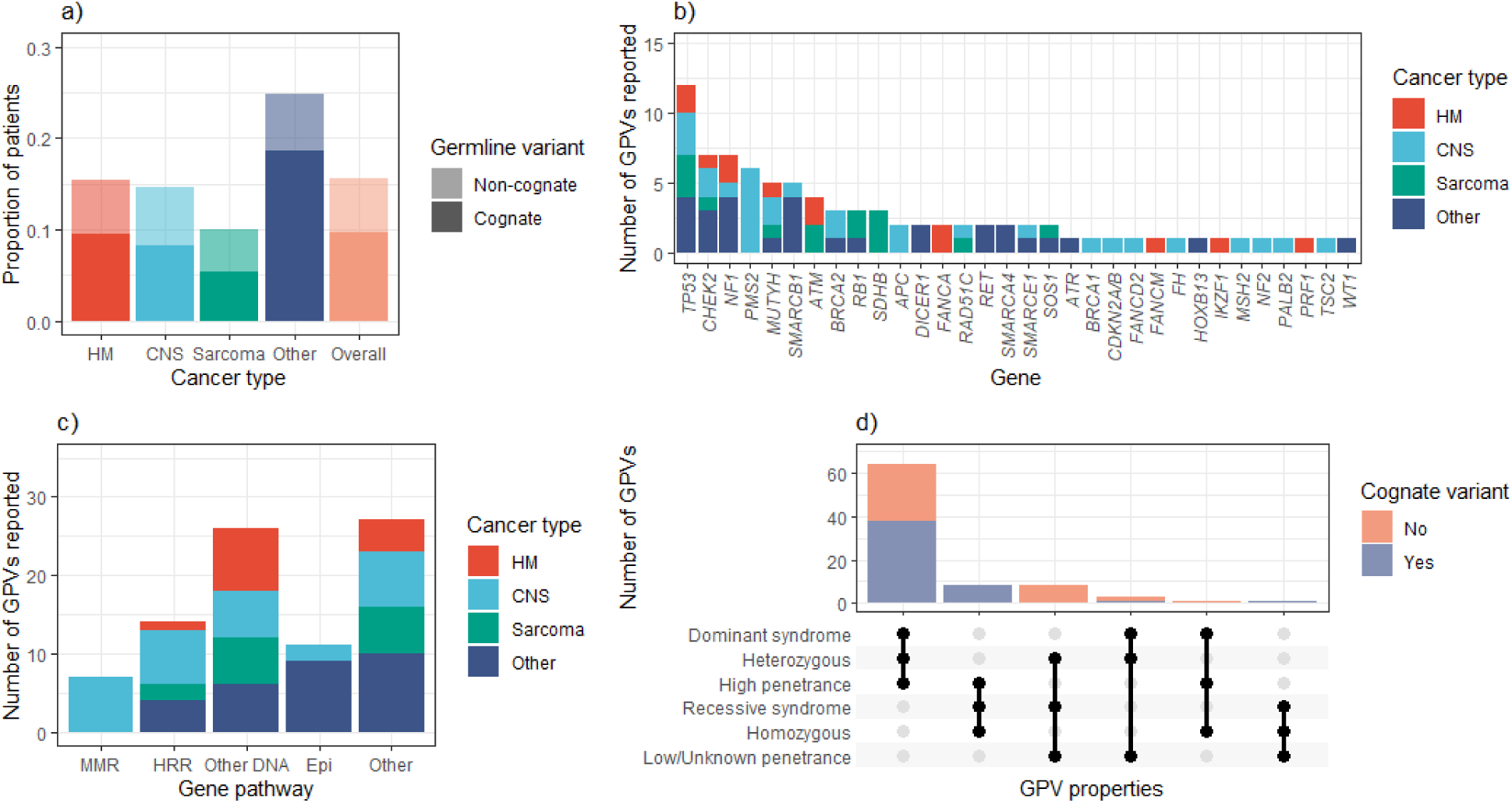
Overview of reported germline findings in PRISM. (a) The rate of GPV varied by cancer type, with most GPV found in genes with a known link to the patient’s cancer (here termed “cognate GPV”). (b) Cancer predisposition genes for which GPV were identified, and (c) gene-cancer type associations emerged when genes were grouped by biological pathway, with the most notable being that germline MMR deficiency variants were exclusively observed in children with CNS tumours. (d) We reported predominantly heterozygous GPV in genes associated with autosomal dominant cancer predisposition conditions of moderate to high penetrance, mostly in keeping with the patient’s cancer type. Abbreviations: HM: haematological malignancy; CNS: central nervous system cancer; MMR: mismatch repair; HRR: homologous recombination repair (dominant acting genes only); Other DNA: other DNA maintenance genes; Epi: epigenetic maintenance and control genes; GPV: germline pathogenic/likely pathogenic variant/s.

### Germline cancer risk findings have high clinical value

The clinical impact of detecting a GPV varies depending on multiple factors. To evaluate the clinical relevance of the GPV in our cohort, we tabulated our reportable germline findings by inheritance pattern, penetrance, and whether there was or was not an established association between the GPV and the patient’s cancer diagnosis (cognate *versus* non-cognate cancer type, respectively). We defined findings of high clinical value as those likely to manifest as a high cancer risk, being high- or moderate-penetrance GPV (relative cancer risk > 2-fold in genes associated with autosomal dominant (AD) cancer conditions, or autosomal recessive (AR) conditions with biallelic GPVClick or tap here to enter text.(Supplementary Table 1)).

We identified high clinical value findings in 14.1% of patients (70/496), comprising 73 of the 85 reported GPV. The GPV of high clinical value were linked to AD conditions (65/73, 89.0%) or were biallelic loss of genes associated with AR conditions (8/73, 11.0%) (Fig. 1d, Supplementary Table 1). Although we did not routinely report heterozygous variants in AR conditions, we did report an additional 8 GPV (9.4%) in a carrier state, in part due to them being informative for family planning, high population carrier frequency (∼1%), or due to emerging evidence of a link to paediatric cancer. The remaining 4 (4.7%) reported GPV were in genes with low or unclear penetrance. Together, these results establish a high burden of penetrant cancer-risk GPV in poor prognosis paediatric cancer. As this burden exceeds the commonly-applied 10% prevalence threshold for cancer risk testing, our findings suggest that cancer-agnostic GPV screening is justified for all children with poor-prognosis cancer.

We observed a high prevalence of clinically important GPV that would be missed by genetic testing targeted to the cancer diagnosis. Of the 85 reported GPV, 37 (43.5%, Fig 1a) were non-cognate (in genes not traditionally associated with the cancer diagnosed in the patient) yet were clinically relevant: 73.0% of these non-cognate GPV were associated with AD conditions of high-to-moderate penetrance. Overall, half (48.1%, 37/77) of the patients with a GPV identified by our study would not have been recommended genetic testing of the appropriate gene based on Australian testing criteria, and 40.0% (30/77) would not have met criteria for clinical cancer genetics referral ^17^. Patients would have similarly not been referred for clinical genetics assessment using guidelines arising from the American Association for Cancer Research Childhood Cancer Predisposition Workshop, where 45.3% (29/65) patients with solid tumours and a GPV identified by our study would not have met criteria^18^.

An immunohistochemistry (IHC) assay for the relevant protein was available as part of routine care for only 36/77 (46.8%) patients with GPV, and performed in only 15/36 (41.7%), highlighting the limitation of IHC as a screening biomarker for germline cancer risk.

In summary, germline WGS in our cohort identified a high prevalence of GPV in C-AYA patients with poor prognosis cancers. Moreover, the reported GPV changed clinical management of the patient and/or their family members in 90.1% of cases. However, the causative role of reported GPV in tumour initiation or progression was uncertain, particularly for findings which were judged to be non-cognate based on current understanding of gene-cancer relationships.

### Paired germline-somatic analysis reveals expected and potentially novel gene-phenotype relationships

Although we identified a high rate of GPV in this cohort, it was unclear whether these variants were causally involved in tumorigenesis, or if they were coincidental. To address this, we reasoned that germline variants that are involved in cancer development would induce changes in the tumour that either indicate positive selection for the germline variant (for example, a second hit ablating a tumour suppressor gene in association with a loss-of-function GPV), or a functional effect (such as high mutational burden and microsatellite instability in the tumours of patients with constitutional mismatch repair deficiency). Accordingly, for each patient in our cohort we reviewed clinical and molecular data for specific correlative features that would indicate a role of the germline variants in cancer development.

We first established the accuracy of our tumour molecular features (TMF). As expected, our molecular measurement of mismatch repair deficiency (MMRD) mutational signatures (MutSig) closely correlated with both germline MMRD, and tumour MMRD as determined by molecular analysis (Fig. 2a). Likewise, our measurement of genome wide loss-of-heterozygosity (gwLOH, a measure of the fraction of the tumour genome that has become haploinsufficient) was significantly associated with loss-of-function of *TP53*, consistent with the established link between loss of *TP53* function and genome instability (Fig. 2b). However, we did not identify any association between germline homologous recombination deficiency (HRD) and relevant TMF, including MutSig linked to HRD in adult cancer (MutSig SBS3, ID6; Fig 2c).

**Figure 2:**
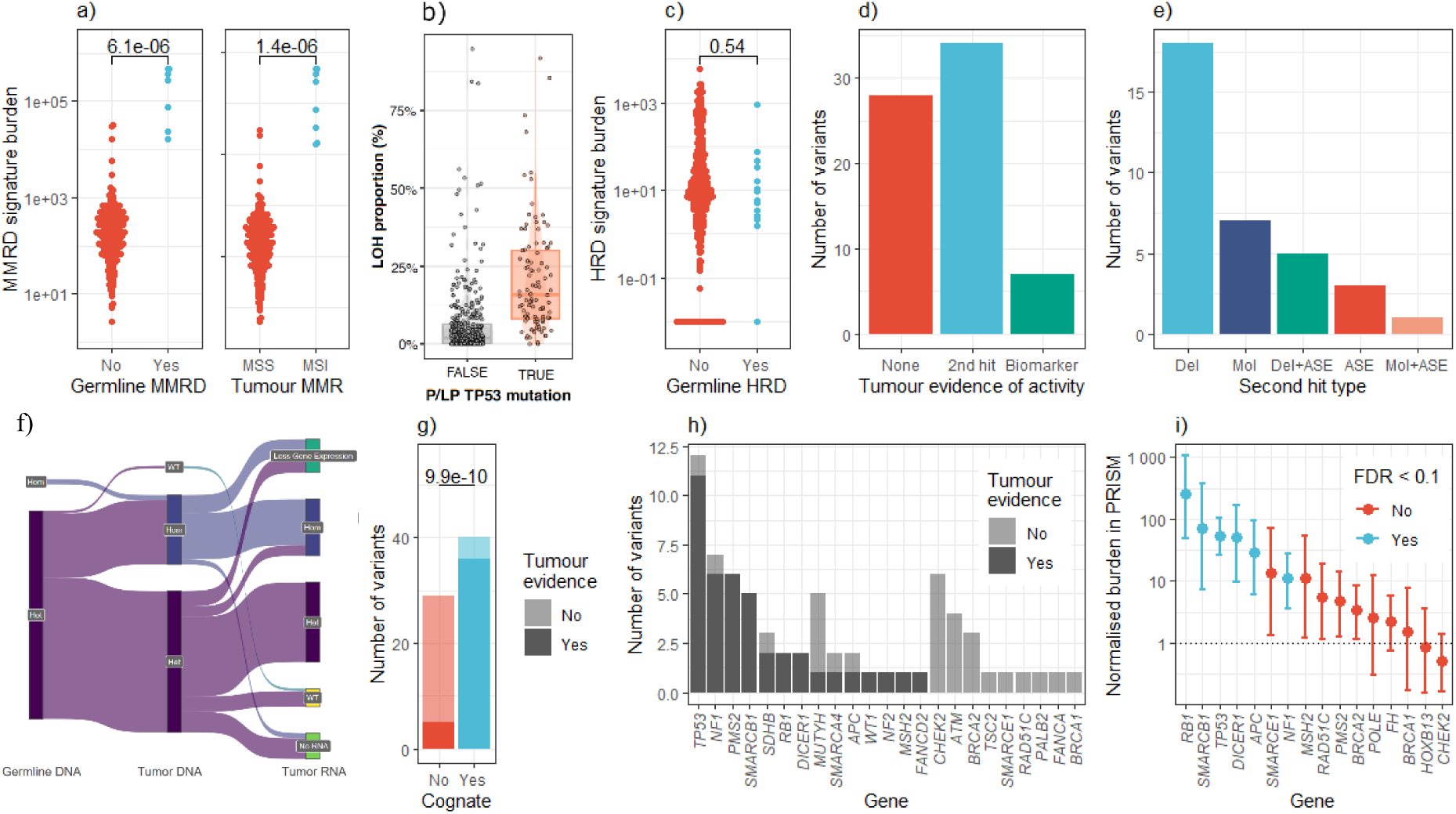
Paired tumour-germline and population analysis implicates key cancer risk genes in the PRISM cohort. (a) Tumour molecular features associated with mismatch repair were significantly higher in patients with either germline or tumour mismatch repair deficiency (p < 6.1×10^-6^). (b) Likewise, higher fractions of the genome in a loss of heterozygosity state were observed in tumours harbouring TP53 loss of function variants. (c) A combined measure of tumour HRD was not significantly associated with GPV in CPG associated with HRD (p = 0.54). (d) Overall, most GPVs had corroborating molecular tumour biomarkers suggesting activity of the risk variant in the tumour, with the most common biomarker being a somatic second hit. (e) Somatic second hits were most commonly DNA deletions; (f) deletions were often associated with loss of expression of the wild-type allele observed by RNA-seq. (g) GPV in participants with cognate cancers were far more likely to have corroborating tumour evidence than GPV in patients with non-cognate cancers, with (h) the rate of supportive TMF varying between genes. (i) A complementary variant burden analysis confirmed that PRISM cases were significantly enriched for functionally significant variation relative to the gnomAD cohort in key paediatric CPG, however, no significant enrichment was observed for any genes in the homologous recombination repair pathway. Abbreviations: Del, deletion; Mol, molecular variant (SNV/small indel); ASE, Allele-specific expression leading to absent wild-type transcript; FDR, False Discovery Rate; P/LP: pathogenic/likely pathogenic. Mutation signature burdens are expressed in arbitrary units. Burden test error bars give approximate bootstrap confidence intervals in PRISM relative to gnomAD, not corrected for multiple testing.

We next evaluated the proportion of GPV for which TMF indicated that the GPV was a key driver of tumorigenesis. This was only possible for 77/85 (90.6%) GPV, as the remaining 8 variants were either in patients with poor-quality tumour data, or were in oncogenes for which correlative tumour changes, such as a second hit, would not be expected. More than half of these 77 GPV (41/77, 53.2%) had associated tumour features supportive of the GPV causal role; the majority (34/41, 82.7%) being a somatic second hit leading to complete loss of a tumour suppressor, and the remainder being MutSig associated with MMRD (Fig. 2d). Second hits were predominantly structural variants leading to loss of the wild-type allele, although we observed multiple types of second hits including small mutations, inversions, and allele-specific expression (ASE) resulting in absent wild-type transcript (Fig. 2e). In four cases, WGS retained both the wild-type and variant allele however, RNA-seq showed ASE for the variant allele confirming the loss of gene function (Fig. 2f). These events were not detected by WGS, highlighting the value of multi-modal molecular analysis to identify somatic second hits. In summary, the majority of germline variants displayed evidence of causal involvement in the development of the cancer through the presence of correlative tumour features, and comprehensive identification of these features required a combination of tumour DNA and RNA analysis.

As expected, TMF supporting the causal activity of a GPV were significantly more common in cognate cancer types: 36/45 variants (80.0%) in participants with cognate cancers had supportive TMF, versus 5/32 variants (15.6%) in the group of non-cognate cancers (Fig. 2g, p = 2.9×10^−8^, Fisher’s exact test). The correlation between GPV and TMF varied between genes, with germline variants in some genes (e.g., in *TP53, NF1,* and *PMS2*) almost always found associated with TMF, and others (e.g., in the homologous recombination repair genes *BRCA1, BRCA2, CHEK2,* and *PALB2*) almost never being associated with corroborating TMF (Fig. 2h). Interestingly, we identified four patients with GPV and cognate cancers for whom we found no relevant TMF, including a meningioma and *SMARCE1* GPV; malignant peripheral nerve sheath tumour and *NF1* GPV; *TP53* and adrenocortical cancer; and *SDHB* and gastrointestinal stromal tumour.

The absence of supportive TMF does not necessarily indicate a CPG is not causally involved in carcinogenesis, as the CPG may not yield diagnostic TMFs, or our molecular assays may be insensitive to detect relevant TMFs. To address this issue and further survey the CPGs causally involved in high-risk paediatric cancer, we undertook an orthogonal analysis to identify CPGs which were statistically over-mutated in our paediatric cancer cases relative to population controls. The CPGs with such an excess burden of mutation in our cancer cohort would be epidemiologically linked to high-risk paediatric cancer. Using the ProxECAT method^19^, we identified a highly significant enrichment of predicted disruptive variation across the 59 CPG that passed data quality filters in both cohorts (functional:proxy odds ratio PRISM:gnomAD 2.44, p = 1.7×10^-7^) (Supplementary Table 2). To confirm the specificity of our test, we established that this enrichment was specific to CPG, and was not observed in a negative control set of genes associated with an unrelated condition (skeletal dysplasia, odds ratio 1.19, p = 0.46). To further investigate the genes underlying the significant enrichment observed across all CPGs, we undertook a gene-wise analysis which identified 6/59 genes (*SMARCB1, RB1, TP53, DICER1, APC,* and *NF1*) significantly enriched for functionally deleterious variation in the PRISM cohort relative to gnomAD, at a false discovery rate of 0.1 or less (Fig. 2i). Notably, no genes associated with HRD were significantly enriched for deleterious variation in PRISM, and a combined gene set test for *BRCA1, BRCA2,* and *PALB2* was non-significant (p = 0.25 after Holm multiple test correction). This finding, combined with the lack of corroborating somatic evidence for HRD (Fig. 2h), suggests that germline loss-of-function in HRD genes is not a major driver of cancer initiation in our cohort. However, this finding is statistical in nature and does not exclude HRD gene variation being causal in cancer types not well-represented in our cohort.

### Tumour molecular phenotype is a valuable resource for germline interpretation

We next examined whether tumour features enhanced the detection and interpretation of germline cancer risk findings. In 11 of 77 patients with GPV (14.3%), tumour features aided germline findings (Fig. 3, Supplementary Table 3.). We grouped these 11 cases into three categories based on how TMF influenced GPV interpretation: cases where the TMF triggered manual review of germline WGS data that led to the identification of a previously missed GPV (*Group A*); cases where TMF directly contributed to variant classification (G*roup B*); and cases where TMF suggested a novel causal role of the GPV in cancer development (*Group C*).

**Figure 3.**
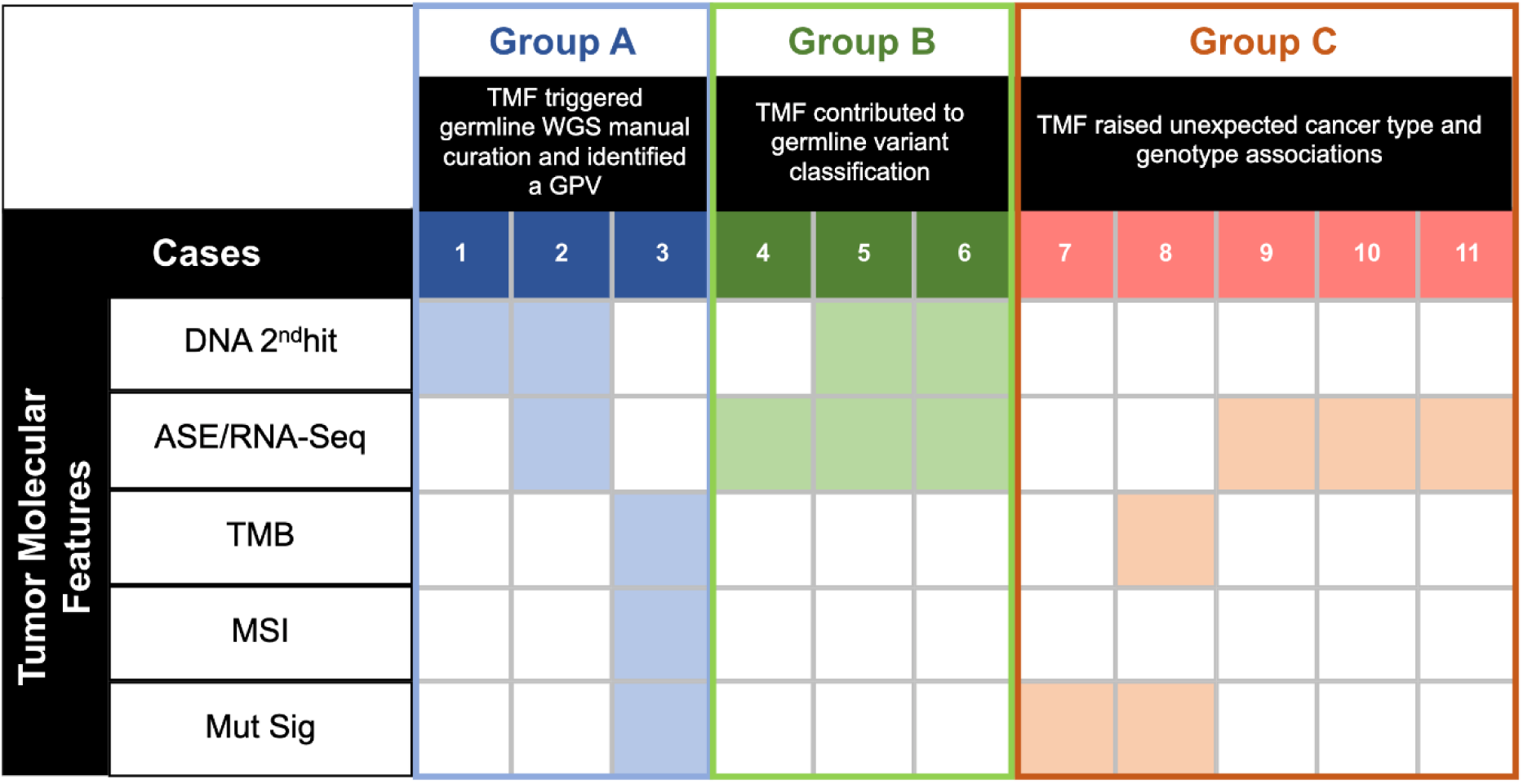
Concordance and direct impact of paired tumour-germline analysis and interpretation. Eleven cases demonstrating the utility of tumour molecular features (TMF) for GPV interpretation categorised by the following: (1) *Group A*, TMF triggered manual curation of WGS germline and identification of GPV; (2) *Group B*, TMF contributes to the classification of GPV (from VUS); and (3) *Group C*, TMF raised potential unexpected association between cancer type and gene-phenotype. Abbreviations: 2ndhit: second hit; ASE: allele specific expression from RNA-Seq; TMB: tumour mutational burden; MSI: microsatellite instability; MutSig: mutational signature; GPV: germline pathogenic/likely pathogenic variant/s.

*Group A*. In three cases, TMF prompted further germline analyses that led to the identification of a reportable GPV missed by the initial molecular analysis (Supplementary Table 3). For example, in one case of SHH subtype medulloblastoma (Case 3, Supplementary Fig. 2a), the TMF, including tumour mutational burden (TMB), microsatellite instability (MSI) and MMRD MutSig, were suggestive of constitutional MMRD (CMMRD), despite only one *PMS2* splicing variant being identified by our pipeline. This led to a recommendation for the clinical testing of *PMS2*, including CNV and SV analysis. As the participant was deceased at the time of referral, clinical *PMS2* testing was performed in the participant’s parents. This confirmed the splicing variant in one parent and identified an additional indel variant causing a frameshift and premature truncation in the other. Retrospective review of the participant’s WGS data confirmed they were compound heterozygous for both *PMS2* GPVs, consistent with the TMF, and thus molecularly confirming the diagnosis of CMMRD.

*Group B*. In three cases, TMF directly assisted in upgrading a variant of uncertain significance (VUS) to a GPV (Fig. 3). For example, a participant with a malignant rhabdoid tumour (Case 4, Supplementary Fig. 2b) was identified to have a heterozygous germline *SMARCB1* tandem duplication of exons 6-7. In isolation this germline finding was curated as a VUS. However, tumour profiling identified copy neutral-LOH (CN-LOH) of chromosome 22, leading to loss of the *SMARCB1* wild-type allele. This confirmed second hit led the PRISM curation team to classify the germline *SMARCB1* tandem duplication as pathogenic, a conclusion which was upheld by subsequent clinical laboratory review.

*Group C*. In this group, TMF supported the potential role of GPV in the development of the specific non-cognate cancer. In one patient with neuroblastoma (case 7), a *BRCA2* GPV was previously identified on a clinical multi-gene panel (undertaken due to a strong family history of cancer suggestive of Li-Fraumeni syndrome) before their enrolment on PRISM. At the time, this *BRCA2* GPV was viewed as a secondary finding. However, tumour analysis performed through PRISM identified a high level of the HRD-associated MutSig 3 in the tumour DNA, raising the possibility that the *BRCA2* GPV is functionally significant. Neuroblastoma has previously been suggested to be associated with *BRCA2* GPV^20^, so this finding supports the extension of *BRCA2*-related cancers to include some neuroblastoma. Although at a cohort level we were unable to demonstrate a significant contribution of GPV involved in HRD, individual cases still demonstrate activity, highlighting the value of our comprehensive approach. Mutational signatures also proved useful in a case with a CNS embryonal tumour and heterozygous *MSH2* GPV (case 8), with MMRD MutSig in addition to high TMB providing evidence of a causal role.

Taken together, the integration of tumour molecular profiling with the analysis of germline WGS informed GPV interpretation in 14.3% (11/77) of participants with GPV reported in the cohort, and led to an 8.5% increase in the GPV detection rate.

### Clinical impact of the reported germline findings

The clinical impact of germline WGS in poor prognosis paediatric cancer patients has not previously been reported. We therefore evaluated the clinical outcomes for patients with reportable GPV detected by germline WGS. Germline findings were reported to the treating oncologists for 76/77 (98.7%) patients (return of germline results was declined by one family). Referrals to cancer genetics services for patients with reportable GPV were made at the treating clinician’s discretion. For each patient with a reported GPV, surveys assessing clinical utility of the germline finding were circulated to recruiting oncologists and genetic clinicians associated with the study. The survey response rate was 32.5% (25/76) for paediatric oncologists, and 97.4% (74/76) for genetic clinicians (Supplementary Methods).

Oncologists felt that the germline findings enhanced their understanding of their patient’s cancer type in 56% (14/25) of cases. The reported GPV directly informed clinical care in four (16.0%) cases by impacting the immediate cancer treatment in a patient with MMRD-related high-grade glioma and recommendation of immunotherapy; informing the risk management in two patients for whom their surveillance plan was modified upon completion of treatment; and contributing to cancer risk stratification in one patient with treatment-related acute myeloid leukaemia and *TP53* GPV.

Responses from genetic clinicians indicated that WGS was more sensitive to detect clinically actionable GPV across a broad range of CPG than standard clinical testing pathways: more than half the patients (39/74, 52.7%) with reportable GPV detected through PRISM not been previously identified through clinical care. As expected, the patients who had GPV identified by clinical care tended to be associated with conditions with features identifiable during childhood. For example, all six patients with NF1 and five of six patients with CMMRD were known prior to PRISM results disclosure (Fig. 4a). For three patients, the GPV detected by PRISM had been identified previously in extended family members and thus revealed obligate carrier status for the intervening relatives, highlighting an important consideration for mainstreamed genomic consent. Five patients with reportable GPV first identified by PRISM (12.8%, 5/39) had undergone prior cancer genetics assessment, two of whom had a clinical history suggestive of the detected GPV but were not offered genetic testing, either due to not meeting clinical testing criteria, or as clinical testing was not available for the relevant gene.

**Figure 4.**
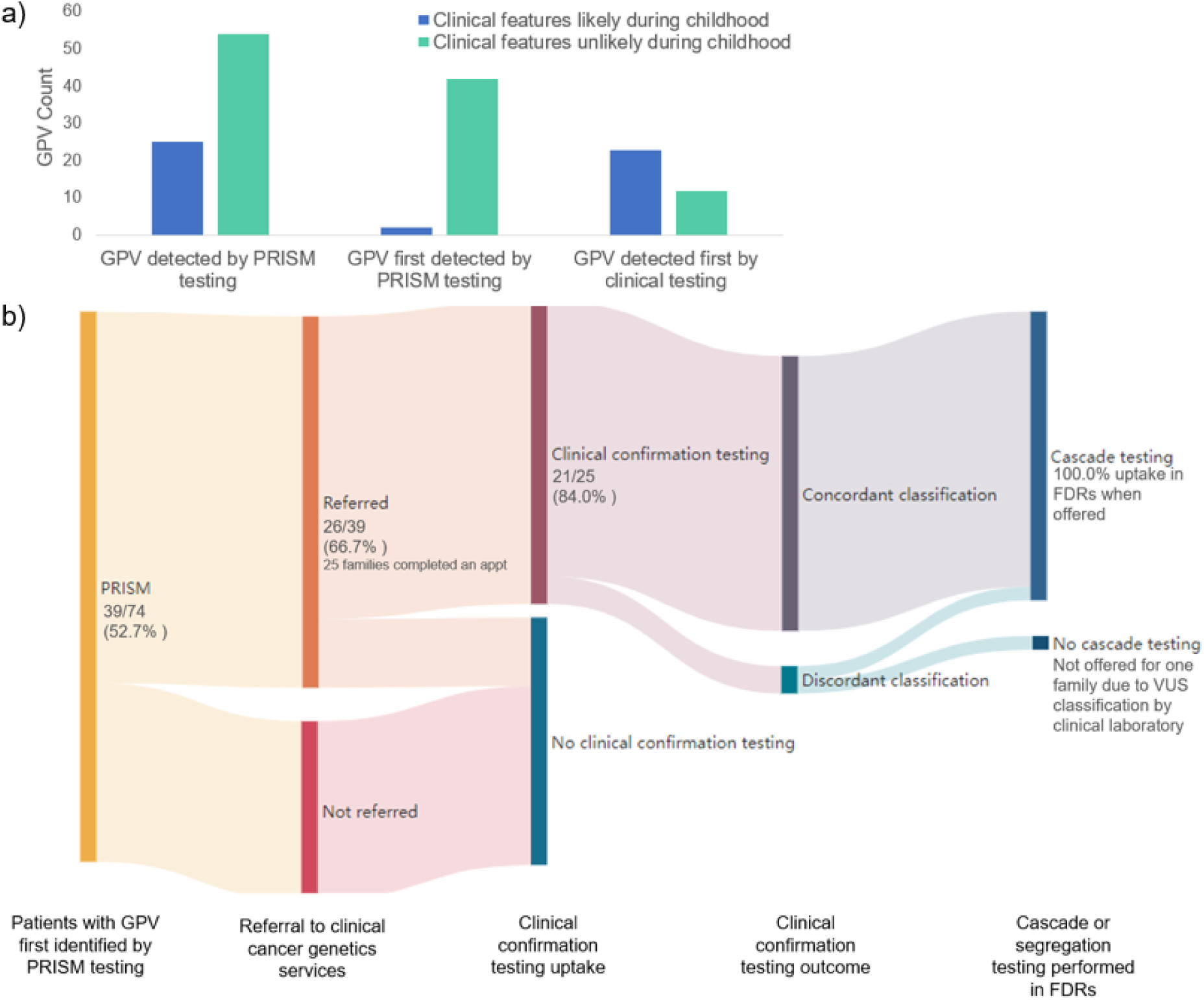
Clinical translation of reported GPV identified by PRISM. (a) The PRISM model of multi-modal CPG screening had a higher GPV yield than clinical testing, especially for GPV associated with conditions that are unlikely to present with clinical features during childhood as compared to those first detected by standard clinical pathways. (b) PRISM findings led to genetics referral and subsequent confirmatory testing in 66.7% of cases, which was concordant with PRISM in 90.4% of cases. Cascade testing in first-degree relatives occurred in all families for whom it was recommended. Abbreviations: FDR, first-degree relative; GPV, germline pathogenic/likely pathogenic variant/s; VUS, variant of uncertain significance; WGS, whole-genome sequencing.

Referrals were received by clinical cancer genetics services for two-thirds (26/39, 66.7%) of patients requiring consultation regarding GPV first identified by PRISM analysis (Fig. 4b, Supplementary Fig. 3). All but one family (25/26, 96%) completed an appointment and there was a high uptake of clinical confirmation testing (21/25, 84.0%). Concordance between research and clinical testing was high, with only two patients having discordant variant classification: in one case a *WT1* variant found in a Wilms tumour patient was assigned a likely pathogenic classification by PRISM due to somatic loss of the *WT1* wild-type allele (case 6, Supplementary Table 3), the clinical laboratory did not incorporate somatic findings into its curation and assigned the variant a VUS classification; in the second case a *RB1* variant in a patient without retinoblastoma was assigned likely pathogenic by PRISM, but was classified as VUS by the clinical laboratory as the variant had not been reported previously in any individual with *RB1*-related disease.

Cascade testing occurred in all first-degree relatives for whom it was recommended (20 families), confirming parental lineage for 73.9% (17/23) of the GPV, with 21.7% (5/23) identified as *de novo* (including three *SMARCB1,* one *TP53* and one *MSH2*). Inheritance was unable to be determined for one family as relatives were unavailable for testing. Australian clinical management guidelines recommend a change in cancer risk management for the relevant parent in eleven (11/17, 64.7%) families where parental lineage was confirmed. Overall, cascade testing occurred in the first-degree relatives of 51.3% (20/39) of participants with reportable GPV first identified by PRISM. Among families who had completed an appointment, genetics clinicians reported discussing reproductive options prompted by PRISM results with parents of seven patients (28.0%, 7/25). The predominant reason provided for not discussing reproductive options was that the parents were not planning to have further children (52.0%, 13/25).

Our results demonstrate the high willingness among families of children with poor prognosis cancers, identified to have GPV through a precision medicine program, to undergo clinical confirmation and cascade testing, even in non-cognate findings where the causal relationship with their child’s cancer is unclear. This indicates there is utility in reporting these findings in this setting, especially for the management of at-risk relatives.

## Discussion

Here we present the germline findings from the first 496 consecutively enrolled patients on PRISM, a national study offering comprehensive germline WGS analysis, integrated with tumour WGS, RNA-Seq, and DNA methylation profiling, to C-AYA with poor-prognosis cancers. We demonstrate a high rate of genetic cancer predisposition, with 15.6% of paediatric patients found to harbor reportable GPV. A substantial proportion of these variants were associated with highly penetrant and AD cancer risk, even for patients with variants in genes not known to be associated with risk of their cancer type. Critically, integrating tumour molecular profiling with the analysis and interpretation of germline WGS increased the GPV detection rate relative to germline-only testing, and informed the underlying causal relationship between the GPV and the patient’s cancer. To our knowledge, this is the first prospective, multi-institution, national study demonstrating the feasibility and clinical utility of integrating germline WGS with somatic tumour profiling to detect germline cancer predisposition in children.

Our study confirms that GPV is prevalent in poor-prognosis paediatric cancers and demonstrates the clinical value of a tumour agnostic screening approach for GPV diagnosis. Consistent with previous reports^1–9^, we found a GPV rate exceeding 10% across all tumour groups, with 13.9% of children carrying clinically actionable findings that confer a moderate to high risk of cancer. Under current Australian clinical genetic testing criteria, this suggests that germline testing is warranted as standard-of-care for all poor-prognosis paediatric cancer patients. Crucially, almost half of all patients identified with a GPV in our study did not meet gene-specific clinical testing criteria, supporting the use of tumour agnostic GPV screening in these patients. We found no clinical variables significantly associated with the presence of GPV, further highlighting the limitations of a guidelines-based, phenotype-driven testing approach in the paediatric setting.

A consequence of broader testing is the increased likelihood of identifying GPV in cancer types that are not within the established phenotypic spectrum of a CPG (which we termed non-cognate cancers). Almost half (43.7%) of GPV detected in our cohort were in patients with non-cognate cancers, raising questions about the potential role of these variants in oncogenesis. The ability to interrogate tumour molecular profiling data in tandem with germline WGS analysis enabled us to evaluate the causal role of novel GPVs prior to variant reporting and proved an important strength of the ZERO multi-omics platform (Fig. 5). Crucially, 15.6% of GPV found in patients with non-cognate cancers had tumour molecular evidence supporting their causal role. This is approximately three times the rate previously reported in non-cognate paediatric cancers among the MSK-IMPACT cohort (5.5%)^21^. However, MSK-IMPACT considered only somatic second hits in the DNA sequence, and our platform incorporated mutational signature and RNA-seq analysis that provided critical corroborative data, potentially accounting for the discrepancy (Fig. 3). Our findings suggest that the paediatric cancer spectrum of CPGs is broader than currently applied in clinical practice, and that cohort studies such as ours that integrate tumour molecular profiling into the assessment of germline variation will expand the CPGs of suspicion for paediatric cancer.

**Figure 5.**
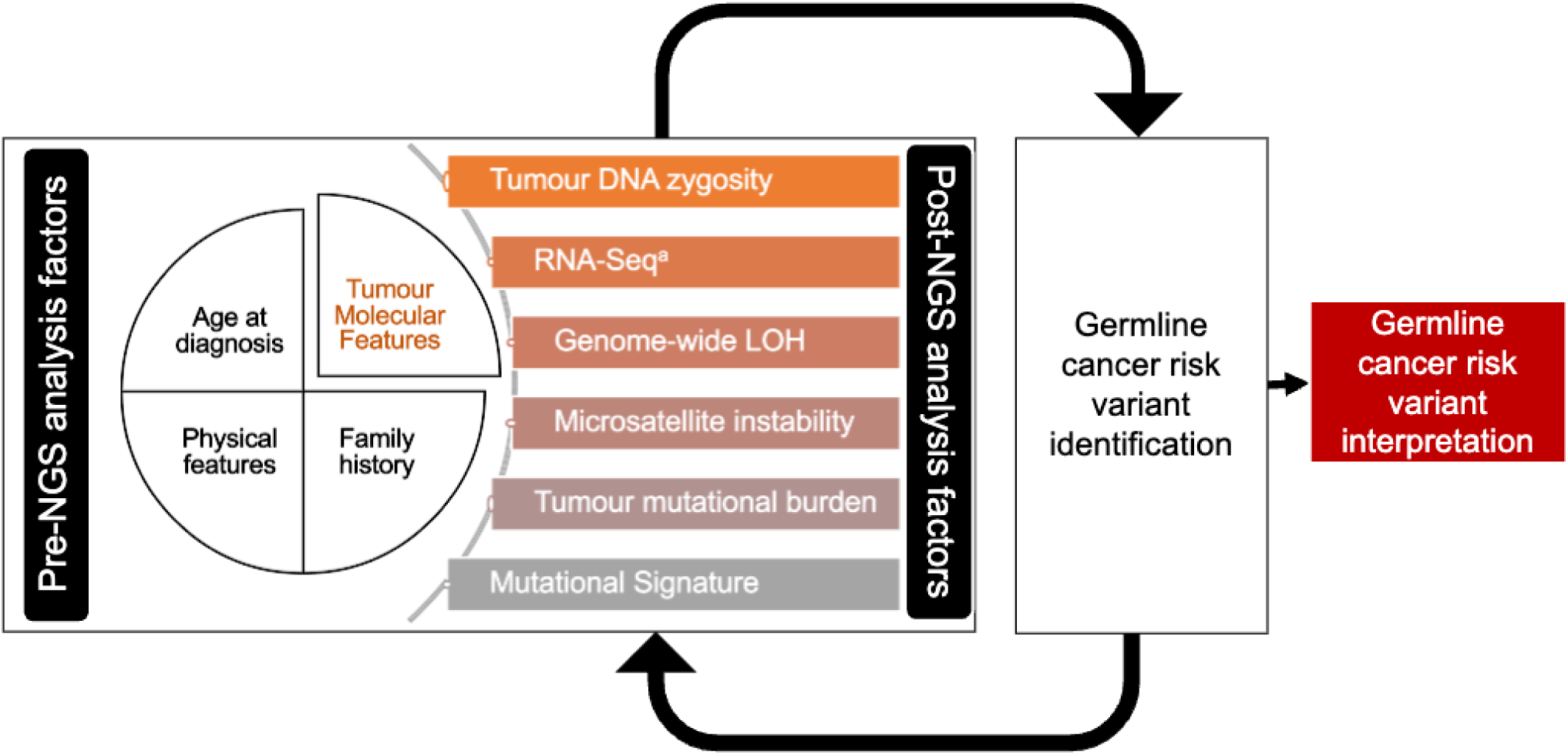
Factors to consider in germline cancer risk variant interpretation. Abbreviations: NGS: next generation sequencing; LOH: loss of heterozygosity. ^a^Refers to RNA-Seq and allele-specific expression.

The availability of comprehensive tumour molecular profiling also facilitated the interpretation of germline WGS (Fig. 3). This was most evident for the six patients for whom tumour data enabled a definitive germline diagnosis, either by prompting manual curation of WGS data (Fig. 3, Group A) or upgrading variant classification from VUS to likely pathogenic (Fig. 3, Group B). In Group A, tumour molecular profiling compensated for known limitations of germline WGS to detect specific variants, and the challenge of keeping germline gene lists updated as novel CPG are described. For Group B variant reclassification was predominantly facilitated by tumour RNASeq and LOH, underscoring the utility of tumour RNA-Seq in addition to tumour DNA sequencing as complementary assays to germline WGS.

Our combined analytic approach provides new insights into the drivers of poor prognosis paediatric cancer. Combining our somatic second hit analysis with a variation burden test, we confirmed the well-established association between childhood cancer and GPV in key CPG such as *TP53*, *RB1*, and members of the SWI-SNF complex (Fig. 2h-i). Notably, however, we could not find evidence supporting the involvement of HRD in cancer development at the cohort level. However, this finding does not exclude HRD being causative in specific paediatric cancers that are not highly represented in our study cohort.

Our results indicate that a screening approach to GPV detection using WGS in paediatric cancer is clinically impactful. More than half of the patients identified with GPV by WGS had not been identified through clinical pathways, despite at least five of these patients undergoing a prior genetics assessment. The high uptake of clinical confirmation and cascade testing in the first-degree relatives (51.3%) is much higher than the 21.0% rate previously reported^7^. Most importantly, the potential for confirmation and cascade testing to result in changes in risk management in almost two-thirds of families highlights that a tumour-agnostic WGS approach in C-AYA with poor prognosis cancers has utility that extends beyond the cancer-affected patient. Further research is necessary to evaluate family members’ perspectives of utility and long-term cost-effectiveness analysis regarding the targeting of cancer risk mitigation strategies to the appropriate relatives of children and AYA diagnosed with cancer. That one-third of patients with GPV had not been referred to cancer genetics services suggests there are barriers to uptake other than variant discovery.

This study is limited by its focus on poor prognosis cancers, and it is unclear whether our findings can be extrapolated to all C-AYA diagnosed with cancer. Furthermore, GPV rates vary depending on the number of selected CPG and clinical reporting criteria. In this study, we did not report all heterozygous GPV in CPG associated with AR conditions, and our platform was not able to reliably detect somatic mosaicism or imprinting disorders, both important considerations for several paediatric cancer predisposition conditions. The strength of this study is the comprehensive and robust multi-platform approach, which has been previously discussed^22^.

In summary, we have presented a comprehensive analysis of the clinically reportable GPV identified by germline WGS paired with comprehensive tumour molecular profiling in children with poor prognosis cancer. Our tumour-agnostic and multi-modal approach (Fig. 5) yielded a high rate of clinically actionable GPV in C-AYA with poor-prognosis cancers, which supports

broad genetic screening of this patient population as standard-of-care. We observed that paired tumour-germline molecular profiling increased the GPV diagnosis rate and aids in genetic counselling for the families receiving these results. To inform implementation of this approach into routine clinical care, future research must refine and validate the use of TMF for germline interpretation and explore of the utility of these findings from a range of relevant stakeholders, including clinicians, patients, and their family members.

## Methods

### Patients and samples

Patients were treated at participating Australian paediatric oncology centres between 2018-2021. Patients were offered participation in PRISM if they met the following criteria: aged ≤21 years, diagnosed with poor prognosis cancer (defined by overall five-year survival <30%), and had suitable biospecimens available for paired tumour-germline analysis. Patient samples were collected as previously reported ^9^.

### Study oversight

The PRISM clinical trial (NCT03336931) is a clinical study in the ZERO Childhood Cancer Precision Medicine Program, a national initiative in Australia that aims to improve the outcomes of C-AYA with poor prognosis cancers. The study design and baseline characteristics of the first 200 patients enrolled on PRISM have been published previously^9^.

### Germline variant processing and curation

Germline variants were identified in patient whole-genome sequencing data as previously described^9^. We defined a list of CPGs of interest based on literature curation, followed by expert review by genetics clinicians and scientists. Germline variants predicted to affect these CPGs were selected for further curation if they met any of the following criteria: 1) were rare in controls (<1% frequency in the Exome Aggregation Consortium (ExAC) database, gnomAD, MGRB^26^, Exome Sequencing Project (ESP) and 1000 Genomes); 2) were previously annotated as LP/P on the ClinVar database, or 3) were novel loss-of-function variants in tumour suppressor genes. A national variant curation team was established to classify GPV. Curation constituted three consecutive stages: molecular annotation, phenotypic annotation, and integrated tumour-germline analysis.

#### Molecular annotation

The functional consequence of the variant on the protein was evaluated using American College of Medical Genetics (ACMG) and Sherloc guidelines.^23,24^. Variants that were determined to be likely deleterious to protein function proceeded to phenotypic annotation.

#### Phenotypic annotation

The purpose of phenotypic annotation was to complement molecular annotation by considering the variant in its clinical. Phenotypic annotation established if the variant was in a *cognate* cancer type (previously described association between the cancer and GPV) or *non-cognate* cancer type (limited or no evidence of an association between the cancer and GPV). Information about the expected penetrance of different phenotypes associated with the gene was also reviewed (Supplementary Table 2) as well as inheritance pattern for the phenotype of cancer risk.

#### Integrated tumour-germline analysis

A single molecular scientist curated both the germline data and tumour data simultaneously, allowing for the molecular and phenotypic annotations of germline variants to be interpreted simultaneously with the TMF, including TMB, MSI, MutSig scores, and ASE from RNA-seq.. Following manual curation by molecular scientists, each case was presented at a study germline team meeting to determine final GPV classification and reportability.

#### Germline variant reporting pipeline

Following curation, the study germline team, including clinical geneticists and genetic counsellors, assessed germline variants for reportability. Germline variants detected in the list of curated CPG were reported to the referring oncologist if they were either heterozygous GPV in CPG associated with AD conditions or biallelic (homozygous or compound heterozygous) in CPG associated with AR conditions. Heterozygous variants in CPG associated with AR conditions were only reported on a case-by-case basis in certain circumstances, including: (1) if the cancer type was cognate with the AR condition and there was the possibility of a second cryptic GPV unable to be detected with the PRISM WGS pipeline; (2) there were molecular tumour features suggestive of a causal role; or (3) the carrier frequency was considered high in the Australian population and therefore had reproductive implications.. Findings were presented at the national molecular tumour board (MTB) and case discussion was offered to the treating oncologist, to support the interpretation and return of the germline results to participants and their families.

Reportable variants were communicated to the treating team through a Germline Report which included details of the variants, their interpretation in context of the clinical phenotype and provided family history, and relevant management recommendations from the study cancer genetic team, such as a recommendation refer to genetic services or the availability of national risk management guidelines.

### Tumour molecular features

We derived several summary molecular features from tumour sequencing data.

#### Genome-wide loss-of-heterozygosity

Genome-wide loss-of-heterozygosity (GW-LOH) was calculated as the proportion of the tumour genome with an estimated minor allele copy number less than 0.5, based on estimates from PURPLE ^25^.

#### Tumour mutational burden

We identified high-confidence tumour single-nucleotide variants (SNVs) using Sage (https://github.com/hartwigmedical/hmftools/blob/master/sage/README.md), including paediatric-specific hotspots in the calling algorithm. Tumour mutational burden (TMB) was defined as the number of high-confidence variants per megabase of the interrogated genome.

#### Microsatellite instability

Tumour microsatellite instability based on molecular profiling was as reported by PURPLE ^25^.

#### RNA-Seq analysis

Tumour RNA-Seq utilised GATK HaplotypeCaller (v3.6) to identify SNVs. The output was further filtered using the coordinates of the identified GPV and the variant allele frequency (VAF) in the RNA-seq was assessed and compared against the germline and tumour VAF of the variant. ASE was called if the GPV VAF was heterozygous and the RNA-seq VAF was homozygous. Splice site mutations were further assessed by visualising the effects of the splice site mutation using the Integrative Genomics Viewer (IGV) in the RNA-seq to determine of the GPV impacted gene splicing in the tumour. Where possible, predicted splice altering variants were confirmed in RNA sequencing data.

### Variant burden testing

Variant burden testing was performed using the ProxECAT test ^25^. Small genetic variants were interrogated from both the PRISM germline data, and gnomAD v4.0.0, for both cancer genes and a negative control set of skeletal dysmorphia genes derived from the Genomics England PanelApp (Skeletal Dysmorphia panel v4.0) with putative cancer-associated genes removed (see Supplementary Table 2 for gene list). For the ProxECAT test, functional variants were defined as variants with no ClinVar benign/likely benign classification that also had either a ClinVar pathogenic/likely pathogenic classification, or a HIGH functional consequence as judged by Ensembl VEP v111. Proxy variants were defined as variants with no pathogenic/likely pathogenic calls in ClinVar, an Ensembl VEP class of LOW or MODIFIER, all SpliceAI scores 0.05 or less, and no CADD predictions exceeding a Phred-scaled score of 10. Variants overlapping Tier 3 low-quality genome regions ^26^, insertions / deletions 10 bp or larger, or variants present in either PRISM or gnomAD at an allele frequency exceeding 1%, were excluded from the test. ClinVar data were sourced from the Jan 2024 archive. Gene tests employed Benjamini-Hochberg false discovery rate control, and gene set tests were controlled for familywise error by Holm’s procedure ^27,28^

### Clinical follow-up and data collection

A referral to clinical cancer genetics services was recommended for participants with previously unknown reportable GPV, to facilitate clinical confirmation of the research finding and subsequent cascade testing. The timing of these referrals was made at the discretion of the treating clinician. Separate surveys for recruiting oncologists and genetics clinicians involved in the care of each participant were custom designed by PRISM study team members with expertise in clinical cancer genetics, genetic counselling, paediatric oncology, and psychosocial methodologies. Surveys were distributed using the Sydney Local Health District Quality Audit Reporting System for each participant for whom consent was obtained to disclose germline findings. The survey instrument for oncologists assessed prior knowledge of the detected variant and the impact of the finding on the clinical management of the patient. The survey instrument for genetics clinicians assessed prior knowledge of the detected variant, clinical confirmation and cascade testing uptake, and counselling issues.

### Data analysis and statistical methods

Participants’ personal history was used to determine eligibility for clinical testing based on eviQ genetic testing guidelines^29^ or expert consensus. Oncologist-reported baseline family history was collected at enrolment, but due to missing data was not used to determine testing eligibility. Statistical analyses were performed using R 4.3.1.

## Data Availability

All interim data produced in the present study are available upon reasonable request to the authors. Patient molecular data are available through application to the ZERO Data Access Committee, see https://www.zerochildhoodcancer.org.au/clinicians-researchers/for-researchers/data-and-sample-resources

## Acknowledgements

We sincerely thank the patients and their families for participating in this study. We thank the many healthcare professionals for their time in consenting patients and for the collection and coordination of samples and associated clinical data at Sydney Children’s Hospital, The Children’s Hospital at Westmead, John Hunter Children’s Hospital, Queensland Children’s Hospital, Royal Children’s Hospital, Monash Children’s Hospital, Adelaide Women’s & Children’s Hospital and Perth Children’s Hospital. The authors thank the Sydney Children’s Tumour Bank Network for providing samples and related clinical information for this study. We thank the staff of the Zero Childhood Cancer Program team, and the broader Children’s Cancer Institute, for their dedicated work on the development, implementation, and delivery of the PRISM study. We thank national cancer genetics services for their expert input in the development of study methods and care for patients with reportable germline results from PRISM and their families. We thank the staff at the Kinghorn Centre for Clinical Genomics for rapid processing of WGS, staff at the Murdoch Children’s Research Institute/Victorian Comprehensive Genetics Service for rapid processing of RNA sequencing, and staff at the Australian Genome Research Facility for rapid processing of methylation data. We thank the Australian Federal Government Department of Health, the New South Wales State Government and the Australian Cancer Research Foundation for funding to establish infrastructure to support the Zero Childhood Cancer personalized medicine program. We thank the Kids Cancer Alliance, Cancer Therapeutics Cooperative Research Centre, for supporting the development of a personalized medicine program; Tour de Cure for supporting tumour biobank personnel; The Steven Walter Children’s Cancer Foundation and The Hyundai Help 4 Kids Foundation for supporting G.M.M. and P.G.E.; and the Lions Kids Cancer Genome Project, a joint initiative of Lions International Foundation, the Australian Lions Children’s Cancer Research Foundation (ALCCRF), the Garvan Institute of Medical Research, the Children’s Cancer Institute and the Kids Cancer Centre, Sydney Children’s Hospital. Lions International and ALCCRF provided funding to perform WGS and for key personnel, with thanks to J. Collins for project governance and advocacy. We thank the Cure Brain Cancer Foundation for supporting RNA sequencing of patients with brain tumours; the Kids Cancer Project for supporting molecular profiling and molecular and clinical trial personnel; and the University of New South Wales, W. Peters and the Australian Genomics Health Alliance for providing personnel funding support. The New South Wales Ministry of Health-funded Luminesce Alliance provided funding support for computational personnel and infrastructure. The Medical Research Future Fund, the Australian Brain Cancer Mission, the Minderoo Foundation’s Collaborate Against Cancer Initiative and funds raised through the Zero Childhood Cancer Capacity Campaign, a joint initiative of the Children’s Cancer Institute and the Sydney Children’s Hospital Foundation, supported the national clinical trial and associated clinical and research personnel. We thank the Kinghorn Foundation for personnel support. We thank the Cancer Institute of New South Wales (fellowship funding for M.J.C.). We thank the 2018 Priority-Driven Collaborative Cancer Research Scheme, co-funded by Cancer Australia and My Room, for personnel and computational support (grant no. 1165556 awarded to M.J.C.). D.S.Z. is supported by grants from the National Health and Medical Research Council (Synergy Grant #2019056, and Leadership Grant APP2017898) and Cancer Institute New South Wales Program Grant (TPG2037). Zero Childhood Cancer is a joint initiative led by the Children’s Cancer Institute and Sydney Children’s Hospital, Randwick.

## Ethics declarations

Ethics approval for this research was granted by the Sydney Children’s Hospitals Network Human Research Ethics Committee (LNR/14/SCH/497), the Hunter New England Human Research Ethics Committee of the Hunter New England Local Health District (17/02/015/4.06) and the New South Wales Human Research Ethics Committee (HREC/17/HNE/29). Informed consent for each participant was provided by parents/legal guardian for participants under the age of 18 years, and by the participants who were over the age of 18 years.

## ABBREVIATIONS

AD: Autosomal Dominant
AR: Autosomal Recessive
ASE: Allele Specific Expression
C-AYA: Children, Adolescent, and Young Adult
CMMRD: Constitutional Mismatch Repair Disorder
CNS: Central Nervous System
CNV: Copy Number Variant
CPG: Cancer Predisposition Gene/s
Del: Deletion
Epi: Epigenetic maintenance and control genes
FDR: Frist-degree Relative
GPV: Germline pathogenic or likely pathogenic variant/s
gwLOH: Genome wide loss of heterozygosity
Het: Heterozygous
Hom: Homozygous
HRD: Homologous Recombination Repair Deficiency
HRR: Homologous Recombination Repair
LOH: Loss Of Heterozygosity
MINAS: Multilocus Inherited Neoplasia Allele Syndrome
MMR: Mismatched Repair
MMRD: Mistmached Repair Deficiency
Mol: Molecular variant (SNV/small indel)
MSI: Microsatellite Instability
MutSig: Mutational Signature
NGS: Next Generation Sequencing
PRISM: PRecISion Medicine for Children with Cancer
RNASeq: RNA Sequencing
SNV: Single Nucleotide Variation
SV: Structural Variant
TMB: Tumour Mutational Burden
TMF: Tumour Molecular Features
VUS: Variant of Uncertain Significance
WES: Whole Exome Sequencing
WGS: Whole Genome Sequencing

