## Supplemental Methods for "Integrated germline and somatic molecular profiling to detect cancer predisposition has a high clinical impact in poor-prognosis paediatric cancer"

**Survey to Oncology Clinicians regarding PRISM Participants with Reportable Germline Findings**

Participant’s name:

PRISM ID:

Site:

Clinician:

Date of PRISM enrolment:

Germline variant(s) identified:

1. Were you aware that this patient had a pathogenic or likely pathogenic germline variant, prior to the PRISM results?
a. Yes (Go to question 11)
b. No, and it wasn’t suspected
c. No, but I suspected based on the tumour type
d. No, but I suspected based on the family history
e. c and d
f. Suspected based on other reasons - please specify
2. Have you disclosed the PRISM germline results with the patient and/or parents/ guardians?
   a. Yes (go to question 2A)
   b. No, but I plan to share the results with the family in the future (Go to question 5)
   c. No, I do not plan to share the results (go to question 2B)

2A. If yes, how long was it between receiving the germline findings from the PRISM study team and sharing it with the family?

a. 0-30 days
b. 30 - 60 days
c. > 60 days

2B. What are the reasons for not having shared the results with the family? Select all that apply (Go to question 5)

a. Lost contact with the family
b. Patient is deceased
c. The results are not relevant for the family
3. Did you deliver the PRISM germline results together with the PRISM somatic findings?
   a. Yes
   b. No
4. Did the parents receive a copy of the PRISM germline results?
   a. Yes
   b. No
5. In future, do you think that the verbal return of germline results to the families should be separated from the consent for tumour molecular profiling?
   a. Yes – please specify reason
   b. No
   c. I don’t have a preference
6. Did the germline findings contribute to your understanding of your patient’s oncologic condition?
   a. Yes
   b. Not at all
   c. To some extent
   d. To a large extent
7. Did the PRISM germline results modify your perception of your patient’s malignancy risk?
   a. Yes, it contributed to my perception of high-risk malignancy
   b. No, it remained unchanged, it was already considered a high-risk cancer
   c. No, germline findings did not affect risk assessment at all
8. Did the germline findings modify your management of the current malignancy?
   a. Not applicable (patient deceased)
   b. Yes – please specify how
   c. No
9. Did the germline findings modify your plan for surveillance for future malignancies in your patient?
   a. Not applicable (patient deceased)
   b. Yes – please specify how
   c. No
10. Was your patient referred to clinical cancer genetics service?
    a. Yes
    b. No (Go to question 10A)
    c. Unsure
    d. Not yet, but planning to make referral

10A. If no, please specify reason:

a. Unknown
b. Family not interested
c. I don’t think a referral was indicated for my patient
d. Other - please specify

Proceed to Q14

11. If the participant’s germline status was known prior to PRISM results, do you think the information supplied through PRISM was helpful to understand your patient’s condition?
    a. Yes
    b. No
    c. Not at all
    d. To some extent
    e. To a large extent
12. Did the information provided about your patient’s germline finding modify your perception of the malignancy risk stratification?
    a. Yes, it contributed to my perception of high-risk malignancy
    b. No, it remained unchanged, it was already considered a high-risk cancer
    c. No, germline findings did not affect risk assessment
13. Did the germline findings modify your management of the current malignancy?
    a. Not applicable (patient deceased)
    b. Yes – please specify how
    c. No
14. Please provide any additional comments regarding your patient below

________________________________________________________________________________________________________________________________________________________________________________________________________________________________________________________________________________________

**Survey to Genetics Clinicians regarding PRISM Participants with Reportable Germline Findings**

Thank you for helping with additional information on your patient who had a germline pathogenic or likely pathogenic variant reported from the PRISM study.

PRISM ID:

Participant’s name:

Genetic Unit referred to:

Germline variant identified:

Role in this referral:

☐ Doctor

☐ Genetic counsellor

Accompanying information provided

☐ De-identified pedigree indicating: the PRISM participant and which family members have been offered predictive testing by your service

1. Has your service previously been referred paediatric cancer patients?
   a. Yes
   b. No
2. At the time of this survey, have you been referred this patient:
   a. Yes (proceed to question 3)
   b. No, but I am expecting a referral for this patient (end of survey)
   c. No and I am not expecting a referral for this patient (end of survey)
3. What was the priority used to triage the referral for this patient?
   a. Low
   b. Medium
   c. High
   d. Unsure
4. What was the date of referral?
   a. Please specify: _________________________________
5. What was the date of the initial appointment?
   a. Please specify: __________________________________
6. Did you receive a copy of the PRISM germline report along with the referral?
   a. Yes
   b. No
7. Who provided you with a copy of the PRISM germline report?
   a. Referring clinician
   b. PRISM study team
   c. The patient
   d. Other, please specify: __________________________________
8. In your opinion, what level of understanding did the family have regarding why they were referred to genetics?
   a. High
   b. Moderate
   c. Low
   d. No understanding
9. How many appointments has the patient and/or parents had with your service?
   a. 1
   b. 2
   c. 3
   d. 4+
10. Are there additional appointment(s) planned for this patient and/or their parents with your service?
    a. Yes
    b. no
11. How were your consultations with this family delivered? Select all that apply
    a. In-person
    b. Telehealth by videolink
    c. Telehealth by telephone
12. If you have previously seen non-research paediatric cancer patients at your clinic, did this family require
    a. More appointments than a non-research ascertained pediatric clinical referral
    b. The same number of appointments as a non-research ascertained pediatric clinical referral
    c. Less appointments than a non-research ascertained pediatric referral
    d. Our service does not regularly see paediatric patients and their families to provide a compassion
13. Was this germline variant known in the family prior to being identified in the PRISM study?
    a. Yes
    b. No, and it wasn’t suspected
    c. No, but it was suspected based on the tumour type
    d. No, but it was suspected based on the family history
    e. c and d
    f. No, but it was suspected based on other reasons (please specify:_________________________________)
14. Was clinical confirmation of the germline variant offered to the patient and their family?
    a. Yes
    b. No – please specify reason (proceed to question 20)
15. Was the germline variant identified in PRISM clinically confirmed in the participant in an accredited laboratory?
    a. Yes
    b. Yes, it was previously known in the patient
    c. No – please specify reason
16. Have you recommended predictive genetic testing for the germline variant identified in PRISM in other relatives?
    a. Yes
    b. No
17. Was predictive testing offered in relatives? Select all that apply
    a. Yes, first degree relatives
    b. Yes, up to second degree relatives
    c. No relatives have yet attended for predictive testing
    d. No, cascade testing has not be recommended in any relatives
    e. Relatives have seen another service for predictive testing – please specify which service
    f. More predictive testing in the family is planned based on relatives’ results
18. Was predictive testing completed in relatives? Select all that apply
    a. Yes, first degree relatives
    b. Yes, up to second degree relatives
    c. No relatives have yet attended for predictive testing
    d. No, cascade testing has not be recommended in any relatives
    e. Relatives have seen another service for predictive testing – please specify which service
19. Are you aware of any future appointments planned for the family to facilitate cascade testing?
    a. Yes, with our service
    b. Yes, with another service
    c. No
20. Have you recommended additional genetic testing in this family after detailed collection of the personal and family history?
    a. Yes – please specify test type and/or genes
    b. No
21. If known, was the germline variant confirmed to be:
    a. Paternally inherited
    b. Maternally inherited
    c. Mosaic
    d. De novo
    e. Has not yet been determined
    f. Parents were not recommended to have predictive testing
22. Were there published eviQ guidelines available to help guide your management of this family?
    a. Yes
    b. No
    c. Not applicable
23. If a surveillance program was recommended, was the screening subsidised? Tick as many as applied
    a. Yes, by the hospital
    b. Yes, by Medicare
    c. No, it was self-funded
24. In your opinion, please choose which of these counselling issues were apparent to you during your interactions with the patient – select all that apply
    a. Anxiety
    b. Depression
    c. Hopelessness
    d. Grief
    e. Transmission guilt
    f. Fear
    g. Personal control (the patient/their parents felt that genetic testing allowed them to feel in control of their/their children health
    h. Empowerment
    i. Hope
    j. None
    k. Not applicable, patient was too young to discern
    l. Other – please specify
25. In your opinion, please choose which of these counselling issues were apparent to you during your interactions with the patient’s parent(s) – select all that apply
    a. Anxiety
    b. Depression
    c. Hopelessness
    d. Grief
    e. Transmission guilt
    f. Fear
    g. Personal control (the patient/their parents felt that genetic testing allowed them to feel in control of their/their children health
    h. Empowerment
    i. Hope
    j. None
    k. Other – please specify
26. Were reproductive options discussed with the patient and/or their parents? Select all that apply
    a. Yes, IVF with PGD
    b. Yes, prenatal testing
    c. Yes, testing in child soon after birth
    d. Yes, a broad discussion of reproductive options
    e. No, parents are not considering more children
    f. No – please specify reason not discussed
27. Please provide any additional comments regarding your patient below

____________________________________________________________________________________________________________________________________________________________________________________________________________________________________________________________________________
