## Supplemental Table 1 for "Integrated germline and somatic molecular profiling to detect cancer predisposition has a high clinical impact in poor-prognosis paediatric cancer"

**Supplementary Table 2.** Penetrance category assigned to each cancer predisposition gene

| **Gene** | **Penetrance** |
| --- | --- |
| *APC* | High/Medium |
| *ATM* | High/Medium |
| *ATR* | Low/Carrier |
| *BRCA1* | High/Medium |
| *BRCA2* | High/Medium |
| *CDKN2A/CDKN2B* | High/Medium |
| *CHEK2* | High/Medium |
| *DICER1* | High/Medium |
| *FANCA* (biallelic) | High/Medium |
| *FANCA* (monoallelic) | Low/Carrier |
| *FANCD2* (monoallelic) | Low/Carrier |
| *FANCM* (biallelic) | High/Medium |
| *FH* | High/Medium |
| *HOXB13* | High/Medium |
| *IKZF1* | Unknown |
| *MBD4* | Unknown |
| *MSH2* | High/Medium |
| *MUTYH* | Low/Carrier |
| *NF1* | High/Medium |
| *NF2* | High/Medium |
| *PALB2* | High/Medium |
| *PMS2* (biallelic/monoallelic) | High/Medium |
| *PRF1* (biallelic) | Unknown |
| *RAD51C* | High/Medium |
| *RB1* | High/Medium |
| *RET* | High/Medium |
| *SDHB* | High/Medium |
| *SMARCA4* | High/Medium |
| *SMARCB1* | High/Medium |
| *SMARCE1* | High/Medium |
| *SOS1* | Low/Carrier |
| *TP53* | High/Medium |
| *TSC2* | High/Medium |
| *WT1* | High/Medium |
