## Supplemental Table 2 for "Integrated germline and somatic molecular profiling to detect cancer predisposition has a high clinical impact in poor-prognosis paediatric cancer"

**Supplementary Table 2.** List of genes included in variant burden testing

**Gene**

*APC, AXIN2, BAP1, BMPR1A, BRCA1, BRCA2, CDC73, CDH1, CDK4, CDKN1B, CDKN1C, CHEK2, CYLD, DICER1, EGFR, EPCAM, EXT1, EXT2 FH, FLCN, GATA2, HOXB13, LZTR1, MAX, MEN1, MLH1, MSH2, MSH6, NF1, NF2, PALB2, PHOX2B, PMS2, POLE, PRKAR1A, PTCH1, PTEN, RAD51B, RAD51C, RAD51D, RB1, RUNX1, SDHA, SDHAF2, SDHB, SDHC, SDHD, SMAD4, SMARCA4, SMARCB1, SMARCE1, SPRED1, STK11, SUFU, TMEM127, TP53, TSC1, VHL, WT1SD: ACAN, ALPL, ALX4, ANO5, ARCN1, BMP2, CLCN7, COL10A1, COL11A1, COL11A2, COL1A1, COL1A2, COL2A1, COL9A1, COL9A2, COL9A3, DSPP, DVL1, EFTUD2, ENPP1, ERF, FBN1, FGFR1, FLNB, FOXC1, GDF5, GHR, GLI3, HNRNPK, KAT6B, LEMD3, LMNA, LMX1B, LRP5, LTBP3, MNX1, MYH3, NFIX, NIPBL, NOG, NOTCH1, NOTCH2, NPR2, NSD1, PAX3, PIK3R1, PTPN11, PUF60, RAD21, ROR2, RUNX2, SALL1, SALL4, SF3B4, SOX9, TBX3, TBX4, TBX5, TCF12, TCOF1, TGFB2, TGFBR1, TRPS1, TWIST1, WNT1*

Abbreviations: CPGs, cancer predisposition genes;
