## Supplemental Table 3 for "Integrated germline and somatic molecular profiling to detect cancer predisposition has a high clinical impact in poor-prognosis paediatric cancer"

**Supplementary Table 3.** Cases illustrating the utility of integrating tumour and germline analysis for germline variant curation by directly impacting (A) variant detection, (B) variant curation, and (C) the interpretation regarding causation for non-cognate cancers.

|  | **DIAGNOSIS** | **GERMLINE CANCER RISK FINDINGS** | **RELEVANT TUMOUR FINDINGS** | **IMPACT ON GERMLINE INTERPRETATION** |
| --- | --- | --- | --- | --- |
| **Case** | **Group A** | | | |
| 1 | Meningioma  (Patient also has schwannomas) | NM_000268.3(*NF2*):c.115-15G>A | Chr22 loss | Phenotype and somatic monosomy Chr 22 prompted manual search of unfiltered data |
| 2 | Ph-like ALL | NM_006060.6(*IKZF1*):c.40+1G>A | Acquired *IKZF1-IKZF1* inversion  RNA-Seq in keeping | Tumour molecular features prompted manual curation of *IKZF1* in the germline |
| 3 | Medulloblastoma (SHH subtype) | Variant 1: NM_000535.7(PMS2):c.903+1G>C  Variant 2: NM_000535.7(*PMS2*):c.736_741delinsTGTGTGTGAAG (p.Pro246_Pro247delinsCysValTer)  (Compound heterozygous) | TMB 11.49mut/ megab  MutSig SBS6 | Tumour features prompted identification of variant 2 by MLPA |
|  | **Group B** | | | |
| 4 | AML-MRC  (Patient also has FA clinical features, including  increased chromosome breakage) | Variant 1: Deletion *FANCA* exons 12 – 31  Variant 2: NM_000135(*FANCA*):c.1470G>C (p.Gln490His)  (Compound heterozygous) | Tumour RNA-Seq demonstrated read-through of intron 15 (variant 2)  Absence of the wild-type *FANCA* transcript | Shift in classification from VUS to LP of variant 2 |
| 5 | Malignant rhabdoid tumour | *SMARCB1* tandem duplication of exons 6-7 | Chr22 CN-LOH | Shift in classification from VUS to LP |
| 6 | Wilms tumour | NM_024426(*WT1*):c.1051G>A (p.Gly351Arg) | Chr11 CN-LOH | Shift in classification from VUS to LP |
|  | **Group C** | | | |
| 7 | Neuroblastoma | NM_000059.4(*BRCA2*):c.5073dup (p.Trp1692fs) | MutSig SBS3 | Suggests a possible contributing role of the GPV to tumorigenesis |
| 8 | CNS embryonal tumour  (Patient also had a *CHEK2* GPV) | NM_000251(*MSH2*):c.1147C>T (p.Arg383Ter) (heterozygous) | Hypermutated phenotype (45.8 mutations/Mb) and MMRD MutSig | Strongly suggests a contributing role of the GPV to tumorigenesis |
| 9 | Infantile fibrosarcoma | Heterozygous deletion of *SDHB* exon 6 | ASE (RNA 2^nd^ hit) | Suggests a possible contributing role of the GPV to tumorigenesis |
| 10 | Diffuse midline glioma | NM_033084(*FANCD2*):c.2715+1G>A (heterozygous) | ASE (RNA 2^nd^ hit) | Suggests a possible contributing role of the GPV to tumorigenesis |
| 11 | Medullary thyroid cancer  (Patient also had a *RET* and *SOS1* GPV) | NM_001128425.2(*MUTYH*):c.536A>G (p.Tyr179Cys) | ASE (RNA 2^nd^ hit) | Suggests a possible contributing role of the GPV to tumorigenesis |
|  |  | NM_000321.2(*RB1*):c.1222A>C (p.Thr408Pro) | ASE (RNA 2^nd^ hit) | Suggests a possible contributing role of the GPV to tumorigenesis |

Abbreviations: AML-MRC, acute myeloid leukemia with myelodysplasia-related changes; ASE, allele specific expression; Chr, chromosome; DEB, diepoxybutane (DEB); FA, Fanconi anemia; GPV, germline pathogenic/likely pathogenic variant; IHC, immunohistochemistry; LP, likely pathogenic; MLPA, multiplex ligation-dependent probe amplification; MMRD, mismatch repair deficiency; MutSig, mutational signature; P, pathogenic; Ph-like ALL, Philadelphia chromosome–like acute lymphoblastic leukemia; SBS, single base substitutions; SHH, sonic hedgehog; CN-LOH, copy number loss of heterozygosity
