## Supplementary figures and images for "Integrated germline and somatic molecular profiling to detect cancer predisposition has a high clinical impact in poor-prognosis paediatric cancer"

### Supplemental Figure 1

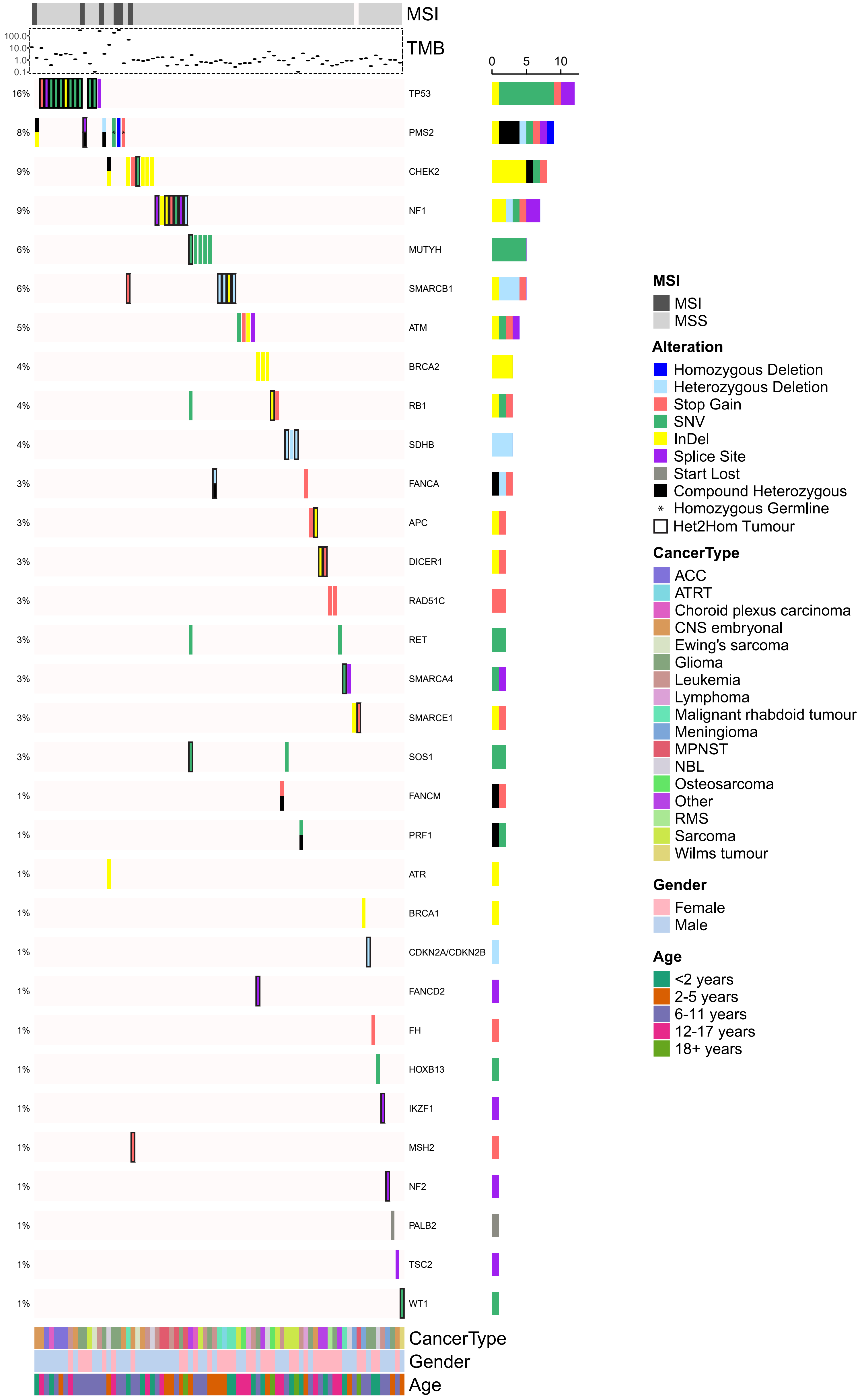
