## Supplemental Figure 2 for "Integrated germline and somatic molecular profiling to detect cancer predisposition has a high clinical impact in poor-prognosis paediatric cancer"

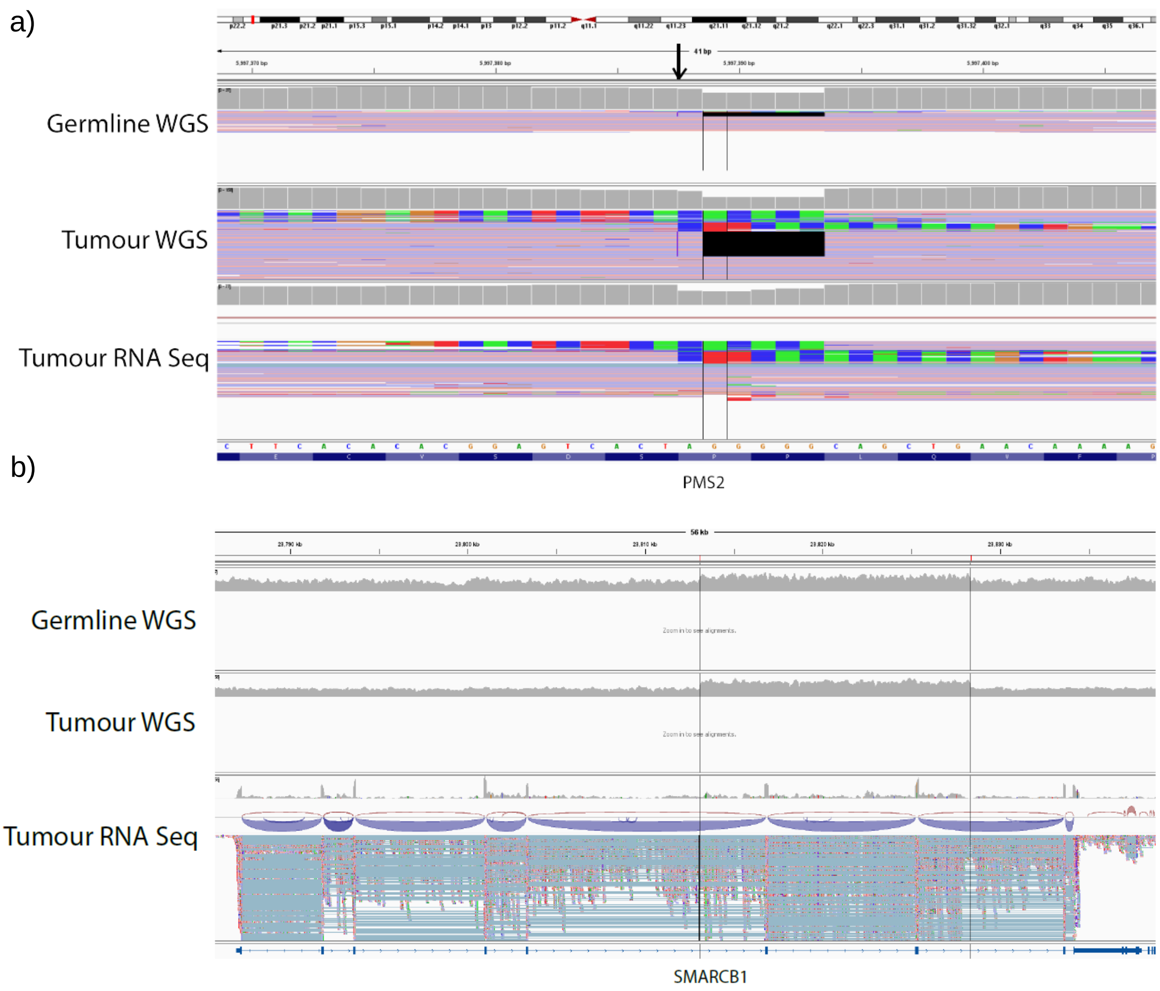


**Supplementary Figure 2. Case examples demonstrating the utility of tumour molecular features to inform germline WGS analysis and interpretation, leading to higher diagnostic yield. (**a) IGV image showing a magnified view of the *PMS2* variant with germline WGS data in the top track, tumour WGS data in the middle track, and tumour RNA sequencing data in the bottom track. The variant inserts 10 base pairs indicated with the purple symbol at the position of the black arrow and deletes 5 base pairs (GGGGG) indicated with the black line. To note, multiple softclip reads are seen in this region that match with this insertion/deletion variant. (b) IGV image showing the entire *SMARCB1* locus with germline WGS data in the top track, tumour WGS data in the middle track, and tumour RNA sequencing data in the bottom track. The two black lines indicate the breakpoint positions for the duplication event that includes exons 6 and 7. Abbreviations: IGV, Integrative Genomics Viewer; WGS, whole-genome sequencing.
