## Supplemental Figure 3 for "Integrated germline and somatic molecular profiling to detect cancer predisposition has a high clinical impact in poor-prognosis paediatric cancer"

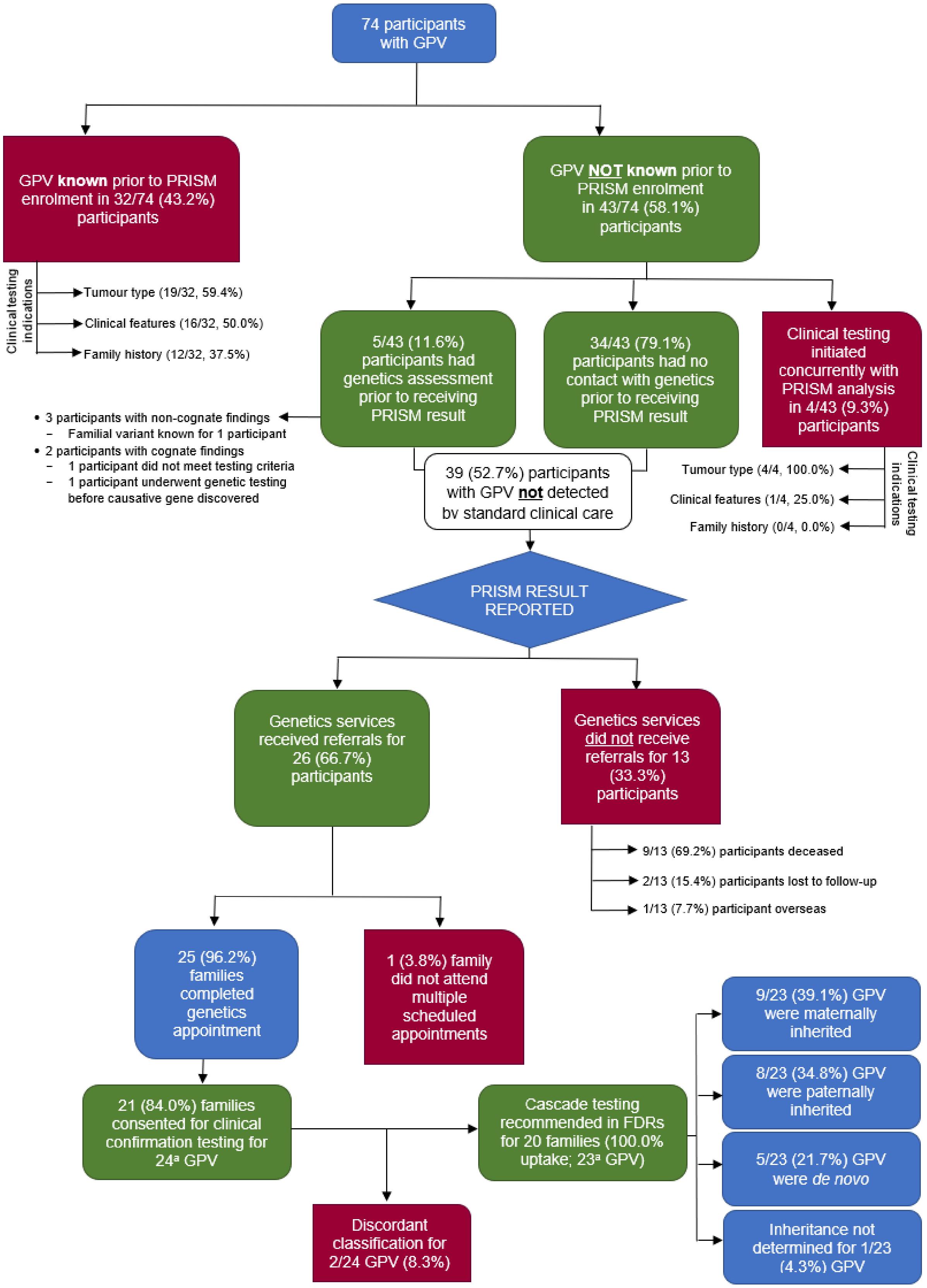


**Supplementary Figure 3. Flowchart of the translation of GPV identified by PRISM research WGS into clinical care.** Uptake rates of clinical confirmation and cascade testing were high among families referred to clinical cancer genetics services. Majority of those participants with GPV detected by PRISM who were not referred to clinical cancer genetics services were deceased at the time of data collection. [a]GPV were counted separately for one patient with compound heterozygous *PMS2* GPV, as each the clinical confirmation and cascade testing outcome pertained to each specific variant. Abbreviations: FDR, first-degree relative; GPV, germline pathogenic/likely pathogenic variant; WGS, whole-genome sequencing.
